# Health Implications of the Nutritional Status of Indian Adults

**DOI:** 10.1101/2020.06.17.20134338

**Authors:** Aalok Ranjan Chaurasia

**Affiliations:** MLC Foundation and ‘Shyam’ Institute, Bhopal, Madhya Pradesh, India

**Keywords:** India, Adults, Nutritional Status, Body Mass Index, BMI, Health Implications

## Abstract

This paper analyses the nutritional status of around 630 thousand Indian adults aged 20-49 years on the basis of their body mass index (BMI) in the context of the implications for the health of adults. The analysis reveals that Indian adults can be divided into 15 mutually exclusive groups on the basis of their nutritional status and individual and household characteristics which has implications not only for health policy and planning but also for productivity and economic growth. The problem of under nourished adults appears to be quite challenging in the poor and the poorest adults of the country living in households without a toilet whereas the problem of over weight and obesity among adults has been found to be largely confined to rich and the richest adults living in the urban areas.

## Introduction

The Body Mass Index (BMI) has been recommended by the World Health Organisation for classifying the relative body weight of adults aged at least 20 years into categories that are associated with increased risk of some non-communicable diseases (WHO, 1995; 2004). As a measure of relative body weight, BMI is easy to obtain. It is an acceptable proxy for thinness and fatness, and has been found to be directly related to health risks and death rates in many studies (Flegal et al, 2005; Willet et al, 2005; Jee at al, 2006; Manson et al, 2007; Klenk et al, 2009; Whitlock et al, 2009; Gonzalez et al, 2010; Flegal et al, 2013; Barell and Samuel, 2014; Bhaskaran et al, 2014; Nuttall, 2015; The Global BMI Mortality Collaboration, 2016; Aune et al, 2016; Klatsky et al, 2017; Bhaskaran et al, 2018). BMI also has a number of advantages for defining the anthropometric characteristics of an adult. It is age and gender independent and is an inexpensive and easy to use tool for screening weight category. It has also been recognised as a useful epidemiological tool for estimating the prevalence of obesity and chronic under-nutrition and the risk of increased morbidity and mortality associated with these nutrition states in adults. Therefore, it has relevance to health policies and programmes in the context of meeting the health care needs, especially, of the adult population. BMI can serve as the basis for segmenting or dividing the adult population in different groups, each having different dominating health and mortality risks, particularly, those associated with the relative weight of the body. It is, however not a diagnostic tool for assessing the health status of an individual.

BMI, essentially, is a proxy indicator for assessing the body build which reflects the physical development of an individual. The ideal measure of the body build is the weight per unit volume of body (Chaurasia and Pattankar, 1979). However, measuring the body volume is quite challenging. The most accurate approach of measuring the body volume is the water replaced by the body while floating. This approach, however, is highly subjective and, therefore, different proxy indexes based on the weight and the height of the individual have been suggested. These indexes can be divided into two categories, relative weight type indexes and power type indexes. A power type index is the ratio of the weight to some power *p* of the height of the individual (Goldbourt and Medalie, 1974). BMI is a power type index with *p*=2. A desirable property of the power type index to serve as the weight per unit volume of the body is that the index should not be correlated or weakly correlated with the height of the individual. In this context, among different power type indexes, the correlation of BMI with the height is found to be the lowest. Recently, an exponential type index has also been suggested as proxy for weight per unit volume of the body (Cidrás, 2015).

In this paper, we analyse the distribution of BMI of almost 630 thousand Indian adults aged 20-49 years and explore how the distribution of BMI is influenced by a host of individual and household level factors. The analysis presents the pan India picture of the nutritional status of adults which has implications for health and mortality. In recent years, a number of studies related to the body weight of adults in relation to their height have been carried out in India (Little et al, 2016; Gajalakshmi et al, 2018; Rai et al, 2018, Rautela et al, 2018; Selvamani and Singh, 2018; Young et al, 2019). These studies, however, do not present the pan-India picture of the nutritional status of adults which has implications for health policy and planning. The analysis follows the segmentation approach which classifies adults aged 20-49 years in a two-dimensional space, one dimension of which is the relative body weight as reflected through BMI while the second dimension is personal and household characteristics of the adult. The segmentation approach divides adults into mutually exclusive groups having distinct personal and household level characteristics and different distribution of BMI. The analysis is relevant to health policy and planning as health and mortality risks in adults have been found to be associated with the relative body weight.

The paper is organised as follows. The next section describes the data used. The analysis is based on the data available through the National Family Health Survey 2015-16 which covered the entire country and which is a part of the Demographic and Health Survey (DHS) Programme being implemented globally. The methodology adopted for analysing data is described in section three while results of the analysis are presented in section four. Section five discusses the findings of the analysis from the perspective of health policy and planning whereas the last section of the paper summarises the main conclusions of the analysis.

## Data

The analysis is based on the data on weight and height of adults aged 20-49 years collected during the National Family Health Survey 2015-16 which is a large-scale household survey conducted in a representative sample of households covering all the states/Union Territories and districts of the country. The survey has two specific goals: a) to generate essential data related to the health and family welfare status of the people needed for policy and programme purposes; and b) to provide information on important emerging health and family welfare issues. The survey covered 601,500 households throughout the country and interviewed 699,686 women aged 15-49 years and 112,122 men aged 15-54 years (International Institute for Population Sciences and ICF, 2017). During the survey, the height and weight of all women aged 15-49 years and all men aged 20-54 years was recorded following the standard protocol of anthropometric measurements. The present analysis is, however, confined to 629754 individuals - 545,704 women aged 20-49 years who were not pregnant at the time of survey and 84,050 men aged 20-49 years as it is argued that BMI is age and gender independent in adults aged at least 20 years. In addition, the survey also collected information about a range of personal and household level characteristics of the adults. The personal level characteristics included, among others, gender, age and the highest level of education attained by the individual. On the other hand, household level characteristics included, among others, residence of the household of the individual, religion and social class of the head of the household of the individual, household wealth index, source of drinking water in the household and sanitation facilities available in the household. This information along with the BMI constituted the basic database for the present analysis.

## Methods

Two classifications have been suggested for classifying adults on the basis of BMI. The first is recommended by the World Health Organization (WHO, 1995) while the second is recommended by a WHO Expert Group, particularly, in the context of Asian populations (WHO Expert Consultation, 2004). The World Health Organization recommends classifying adults into eight categories ranging from severe underweight to type III obesity on the basis of BMI. The WHO Expert Group, on the other hand, has suggested 13 categories of BMI for determining public health and clinical action in the context of Asian populations. The Expert Group observed that the Asian people generally have a higher proportion of body fat than their white counterparts of the same age, sex, and BMI. Moreover, the proportion of Asian people with risk factors for type 2 diabetes and cardiovascular disease is substantial even when the BMI is lower than the WHO cut-off point of 25 kg/m^2^. The Expert Group, therefore, suggested alternative ranges of BMI values which may be associated with low to high risk of type 2 diabetes and cardiovascular disease (WHO Expert Consultation, 2004). The present analysis employs both the classifications to classify Indian adults in terms of their nutritional status.

The BMI of an adult may be influenced by a host of individual and household level characteristics. In order to analyse the influence of these factors on BMI, we have followed the classification modelling approach and applied the classification and regression tree (CRT) method to segments adults into mutually exclusive groups in such a manner that the variation in BMI within a group is as minimum as possible (Brieman et al, 1984). CRT is a nonparametric recursive partitioning method that divides individuals into different mutually exclusive groups in such a way that group homogeneity with respect to the dependent variable is the maximum. The technique sorts individuals into mutually exclusive groups based on that independent variable which causes the most effective split. The process is repeated till either within-group perfect similarity is achieved or the pre-decided stopping criterion is met (Ambalavanan et al, 2006; Lemon et al, 2003). A group in which all adults have the same value of the dependent variable is termed as “pure.” If a group is not “pure,” then the impurity within the group can be measured through different impurity measures including the Gini coefficient of impurity. If the dependent variable is a categorical one, then the method provides the distribution of the dependent variable across adults in each group. If the dependent variable is a continuous one, then the method gives estimates of the arithmetic mean and standard deviation of the dependent variable which reflect the distribution of BMI across adults in each group.

The CRT method of segmentation of adults has many advantages over other classification methods. Perhaps, the most important advantage of the CRT method is that it is inherently non-parametric in approach and so requires no assumption about the underlying distribution of the dependent variable, BMI in the present case. This essentially means that the method can be applied to highly skewed or multi-modal numerical or categorical data with either ordinal or nominal structure also. The method can also be applied to both scale and categorical variables and even the mix of the two. It is, essentially, an automatic “machine learning” method so that relatively little input is required for carrying out the analysis. Last but not the least, CRT output is relatively simple so that its interpretation is straightforward.

In the present analysis, the dependent or the classification variable is the BMI which is a scale variable. On the other hand, the explanatory or the independent variables used in the analysis are: 1) gender of the adult; 2) highest level of the education attained by the adult; 3) residence of the adult; 4) religion of the household of the adult; 5) social class of the household of the adult; 6) source of drinking water in the household; 7) availability of the toilet in the household of the adult; and 8) standard of living of the household as reflected through the household wealth index. All the seven independent variables included in the analysis are categorical variables with two or more than two categories. Actual calculations were carried out using the ‘Tree’ routine of the ‘classify’ module of the Statistical Package for Social Sciences (SPSS) software package.

### BMI in Indian Adults (20-49 years)

On the basis of the WHO 1995 classification (WHO, 1995), almost 18 per cent of Indian adults aged 20-49 years were found to be under-weight as the BMI in these adults was estimated to be less than 18.5 kg/m^2^ according to the National Family Health Survey 2015-16 (Table 1). On the other hand, around 21 per cent of these adults aged 20-49 years were found to be over weight out of which around 16 per cent are pre-obese while about 5 per cent are obese with different degree of obesity. In other words, around 40 per cent of Indian adults aged 20-49 years are vulnerable in the sense that the weight related risk of health and mortality in these adults is relatively high.

**Table 1.**
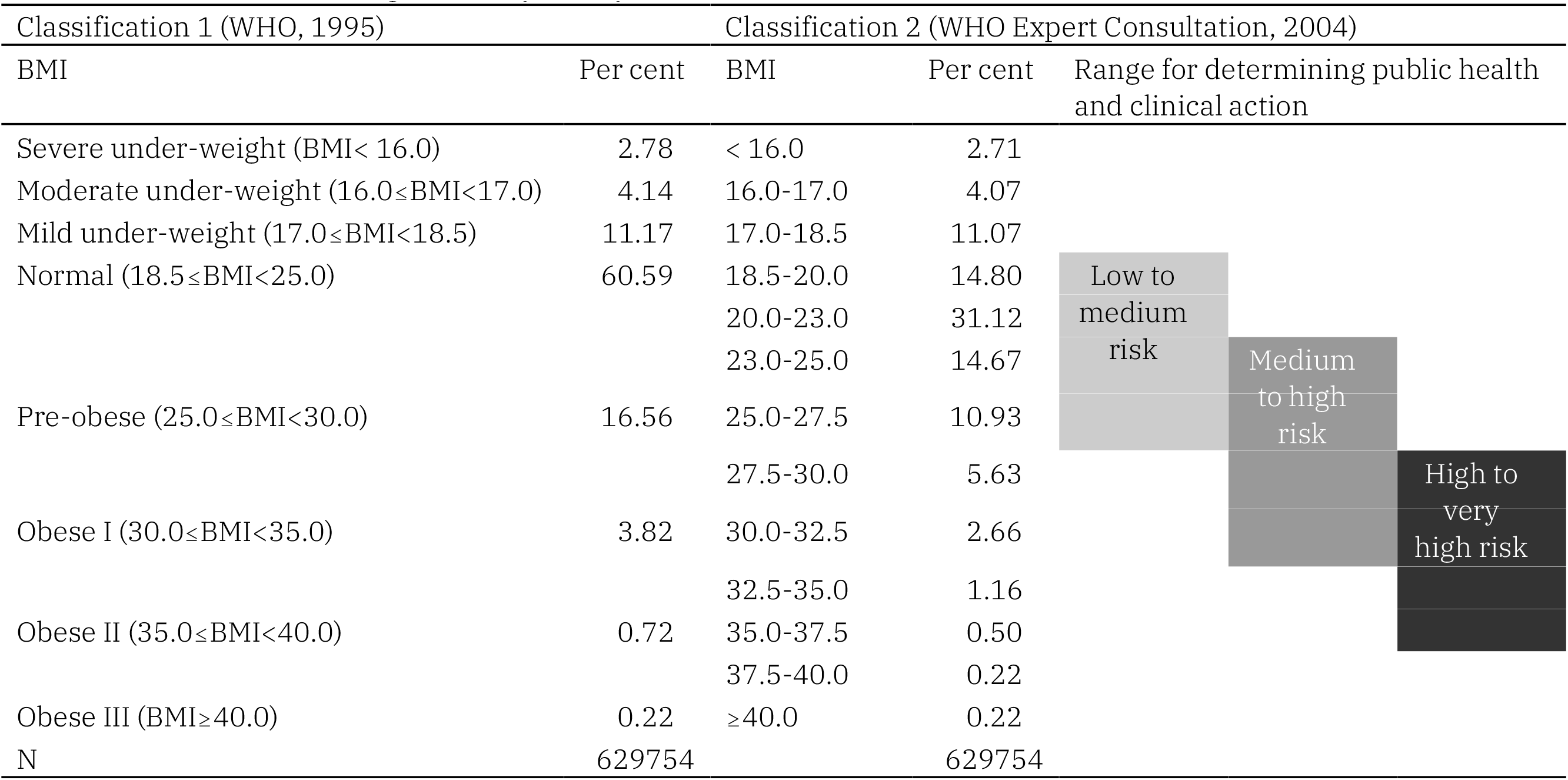
Distribution of Indian adults aged 20-49 years by the level of BMI

On the other hand, following the classification suggested by the WHO Expert Group in the context of Asian populations, almost 72 per cent Indian adults aged 20-49 years appear to be at low to medium risk of type 2 diabetes and cardiovascular disease; more than one third appear to be at medium to high risk; and almost 21 per cent are at high to very high risk of these diseases. This observation, coupled with the fact that almost 18 per cent of Indian adults aged 20-49 years are under-weight suggests that body weight related health and mortality risks in Indian adults are quite substantial and are a major public health challenge.

The distribution of BMI in Indian adults with specific individual and household characteristics is found to be different (Table 2). The mean BMI of rural adults is found to be lower than that of their urban counterparts. There is only a marginal difference in the mean BMI of male as compared to female adults. The level of education attained is also found to be directly related to BMI - the higher the education attained the higher the BMI. Religion and social class also have a strong influence on BMI. The mean BMI is found to be the highest in adults of other religions but the lowest in Hindu adults. Similarly, the mean BMI is found to be relatively the lowest in Scheduled Tribes but relatively the highest in Other Castes. Similarly, BMI appears to be directly related to the standard of living of the adult. The mean BMI is found to be the highest in adults living in households with the richest wealth index quintiles but the lowest in adults living in households with the lowest wealth index quintiles. The gap in the mean BMI of richest adults and the mean BMI of the poorest adults has also been found to be very wide.

**Table 2.** Mean and standard deviation of BMI by individual and household characteristics of Indian adults aged 20-49 years

| Characteristics |  | BMI |  | N |
| --- | --- | --- | --- | --- |
|  |  | Mean | SD |  |
| All |  | 22.19 | 4.18 | 629754 |
| Gender | Male | 22.23 | 3.74 | 84050 |
|  | Female | 22.18 | 4.25 | 545704 |
| Education | No education, preschool | 21.39 | 3.92 | 184570 |
|  | Primary | 21.97 | 4.13 | 92482 |
|  | Secondary | 22.60 | 4.27 | 267724 |
|  | Higher | 22.90 | 4.22 | 83684 |
| Residence | Urban | 23.52 | 4.55 | 188645 |
|  | Rural | 21.62 | 3.88 | 441109 |
| Religion | Hindu | 21.99 | 4.16 | 468087 |
|  | Muslim | 22.74 | 4.45 | 82839 |
|  | Others | 22.80 | 3.87 | 78828 |
| Caste | Scheduled caste | 21.75 | 4.04 | 112051 |
|  | Scheduled tribe | 21.43 | 3.56 | 114856 |
|  | Other Backward Class | 22.17 | 4.23 | 242184 |
|  | None of above | 23.13 | 4.49 | 131034 |
| Drinking water | safe | 22.27 | 4.23 | 564936 |
|  | unsafe | 21.50 | 3.72 | 64818 |
| Toilet | Improved | 23.07 | 4.33 | 370341 |
|  | Unimproved | 22.06 | 3.88 | 31961 |
|  | No toilet | 20.77 | 3.53 | 227452 |
| Living standard | Poorest | 19.89 | 3.00 | 45863 |
|  | Poorer | 20.87 | 3.44 | 204383 |
|  | Middle | 22.49 | 4.09 | 213795 |
|  | Richer | 24.00 | 4.54 | 150268 |
|  | Richest | 24.70 | 4.50 | 15445 |

Source: Author's calculations.

The distribution of BMI is found to be different in the adults living in households having toilet compared to adults living in households not having toilet. The mean BMI is found to be the highest in adults who are living in households having improved toilet facility but the lowest in adults who live in households without toilet. This observation gives credence to the Clean India Mission launched by the Government of India which aims at universal availability of improved toilets throughout the country and elimination of the practice of open defecation. There is, however, little difference in the mean BMI of adults having access to safe drinking water as compared to adults having an unsafe drinking water source.

Results of the segmentation analysis are presented in table 3 while the classification tree is depicted in figure 1. At the first level of classification, 629754 adults are first divided into two groups on the basis of the standard of living - adults with poor and poorest standard of living and other adults. Adults with middle, rich and richest standard of living are further divided into adults with middle standard of living and adults with rich and the richest standard of living. Similarly, adults with poor and the poorest standard of living are further divided on the basis of the availability of toilet in the household. The classification process was continued up to five levels which suggested that Indian adults can be classified into 15 mutually exclusive groups or terminal nodes on the basis of the eight explanatory variables. Every terminal node or group has distinct individual and household characteristics. The most important explanatory variable for classifying adults in terms of BMI is found to be the standard of living followed by the availability of the toilet in the household (Table 4). Relative to the importance of the standard of living, the importance of the availability of the toilet in the household is found to be around 72 per cent whereas that of residence is around 53 per cent. By comparison, the importance of gender, highest level of education attained, source of drinking water, religion and social class has been found to be relatively low.

**Table 3.** Results of the segmentation analysis

| Node | Individual characteristics |  |  |  |  |  | BMI |  | N |
| --- | --- | --- | --- | --- | --- | --- | --- | --- | --- |
| ID | Living standard | Availability of toilet | Residence | Religion | Social class | Highest education attained | Mean | SD |  |
| 0 | All | All | All | All | All | All | 22.19 | 4.18 | 629754 |
| 14 | Poorest | No toilet |  |  |  |  | 19.86 | 3.00 | 42910 |
| 22 | Poor | No toilet |  |  | ST |  | 20.17 | 3.05 | 27911 |
| 21 | Poor | No toilet |  |  | Scheduled Castes,<br>Other Backward Classes,<br>Upper Castes |  | 20.75 | 3.48 | 105345 |
| 11 | Poorest,<br>Poor | Any toilet |  | Hindu |  |  | 21.02 | 3.48 | 40410 |
| 12 | Poorest,<br>Poor | Any toilet |  | Muslim,<br>Others |  |  | 21.67 | 3.38 | 33670 |
| 8 | Middle |  | Urban |  |  |  | 21.85 | 3.96 | 49193 |
| 23 | Middle | Any toilet | Rural | Hindu |  |  | 22.33 | 4.02 | 73925 |
| 24 | Middle | Any toilet | Rural | Muslim,<br>Others |  |  | 22.86 | 3.86 | 34522 |
| 18 | Richer,<br>Richest |  | Rural |  |  | Higher | 22.93 | 4.03 | 16637 |
| 15 | Middle | Any toilet | Urban |  |  |  | 23.03 | 4.32 | 56155 |
| ID | Living standard | Availability of toilet | Residence | Religion | Social class | Highest education attained | Mean | SD |  |
| 26 | Richer, Richest |  | Urban |  | Scheduled Castes, Scheduled Tribes, Other Backward Classes | Higher | 23.51 | 4.27 | 20968 |
| 20 | Richer, Richest |  | Rural |  |  | No education/ Pre-school, Primary, Secondary | 23.90 | 4.45 | 45294 |
| 25 | Richer, Richest |  | Urban |  | Upper Castes | Higher | 24.24 | 4.56 | 16958 |
| 28 | Richer, Richest |  | Urban |  | Scheduled Castes, Scheduled Tribes, Other Backward Classes | No education/ Pre-school, Primary, Secondary | 24.40 | 4.64 | 45584 |
| 27 | Richer, Richest |  | Urban |  | Upper Castes | No education/ Pre-school, Primary, Secondary | 25.03 | 4.88 | 20272 |
| Source: Author's calculations |  |  |  |  |  |  |  |  |  |

**Table 4.** Importance of the independent variables in classifying adults aged 20-49 years on the basis of BMI

| Independent Variable | Importance | Normalized Importance |
| --- | --- | --- |
| Wealth index | 1.902 | 100.0 |
| Type of Toilet | 1.356 | 71.3 |
| Residence | 0.993 | 52.2 |
| Social class | 0.324 | 17.1 |
| Highest educational level attained | 0.228 | 12.0 |
| Religion | 0.063 | 3.3 |
| Drinking water | 0.056 | 2.3 |
| Gender | 0 | 0.0 |

Source: Author's calculations

**Figure 1.**
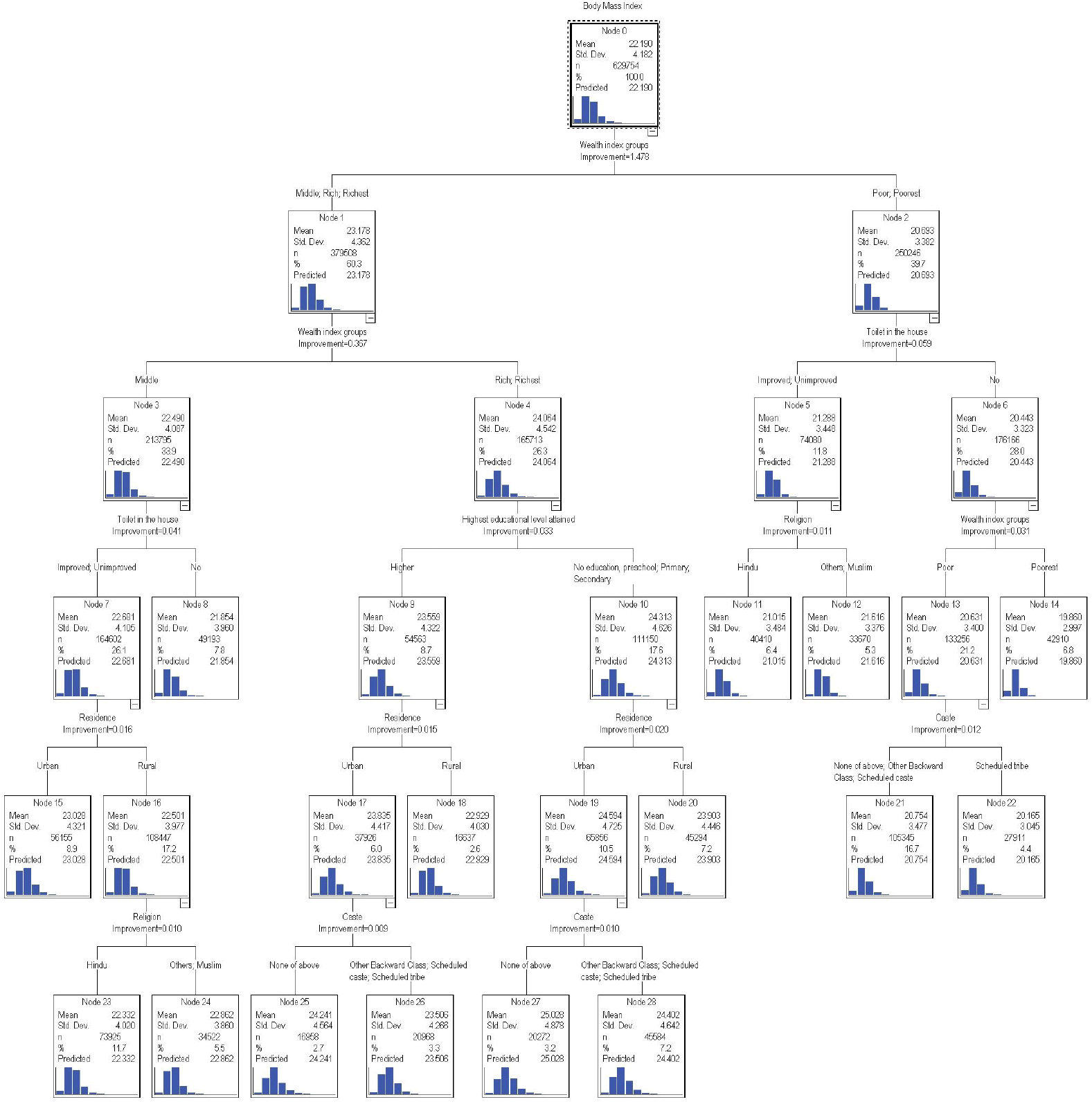
The classification tree.

**Figure 2.**
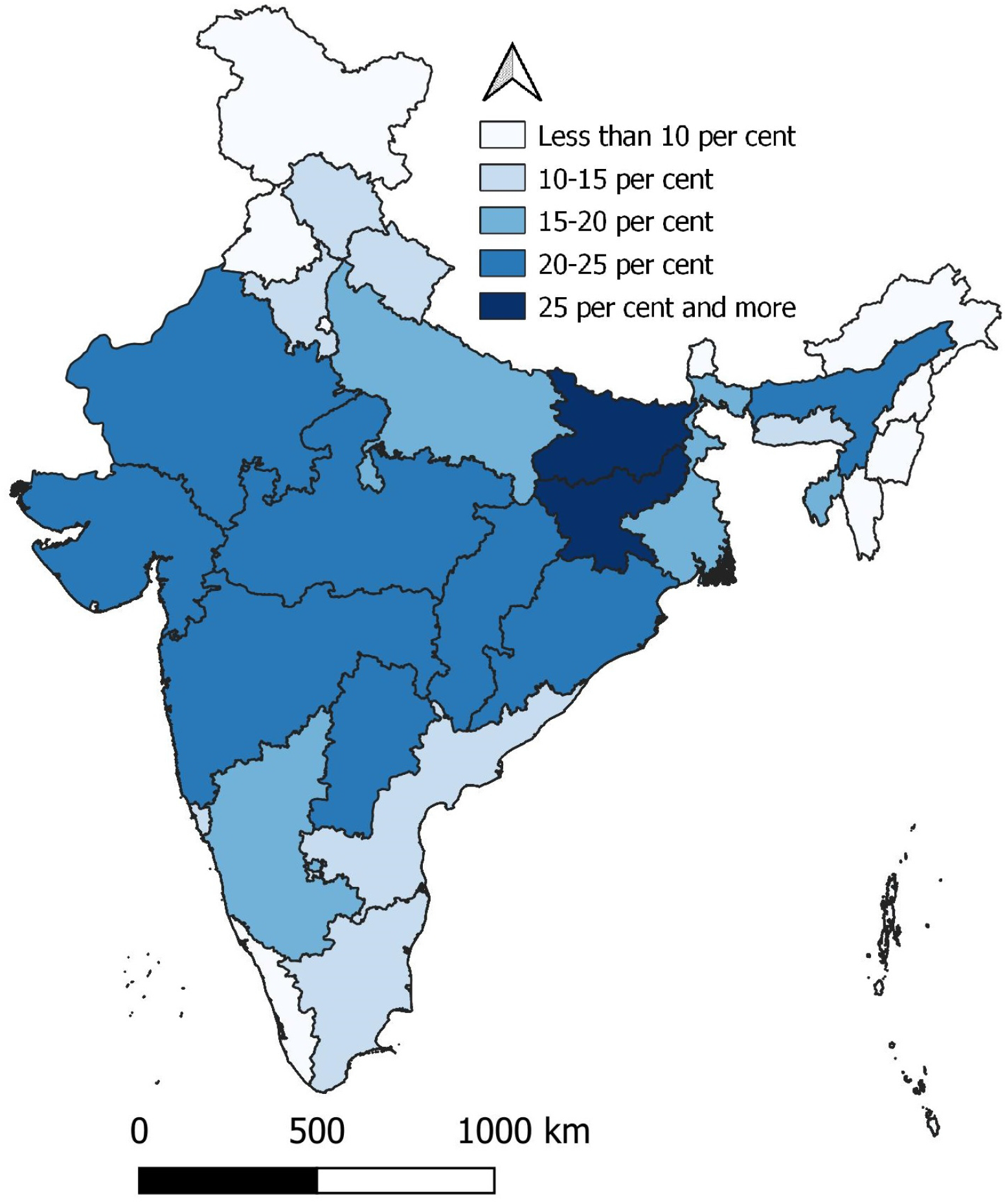
Proportion of adults aged 20-49 years under weight BMI <18.5 Kg/m^2^.

**Figure 3.**
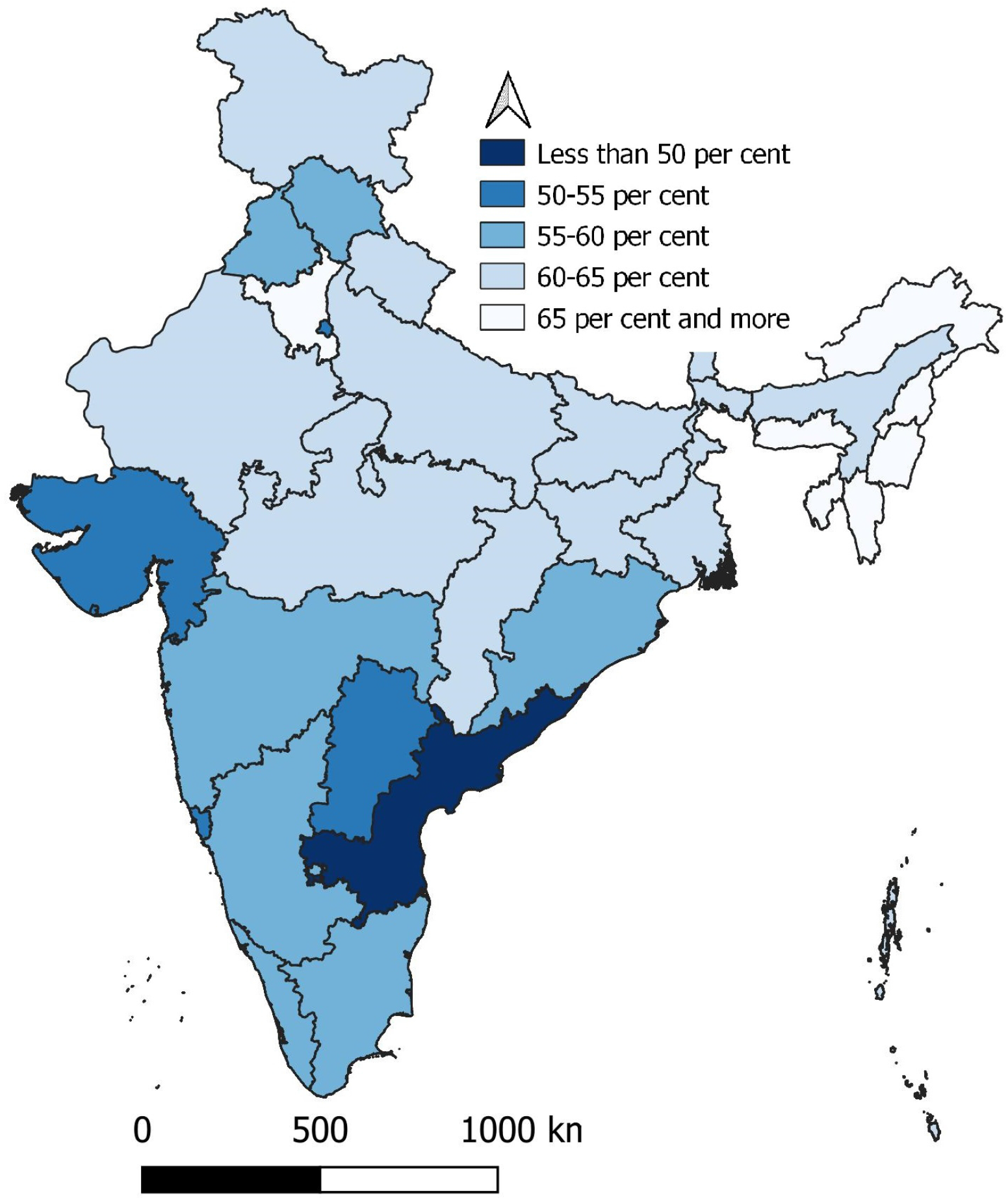
Proportion of adults aged 20-49 years having normal weight 18.5≤BMI<25.0.

**Figure 4.**
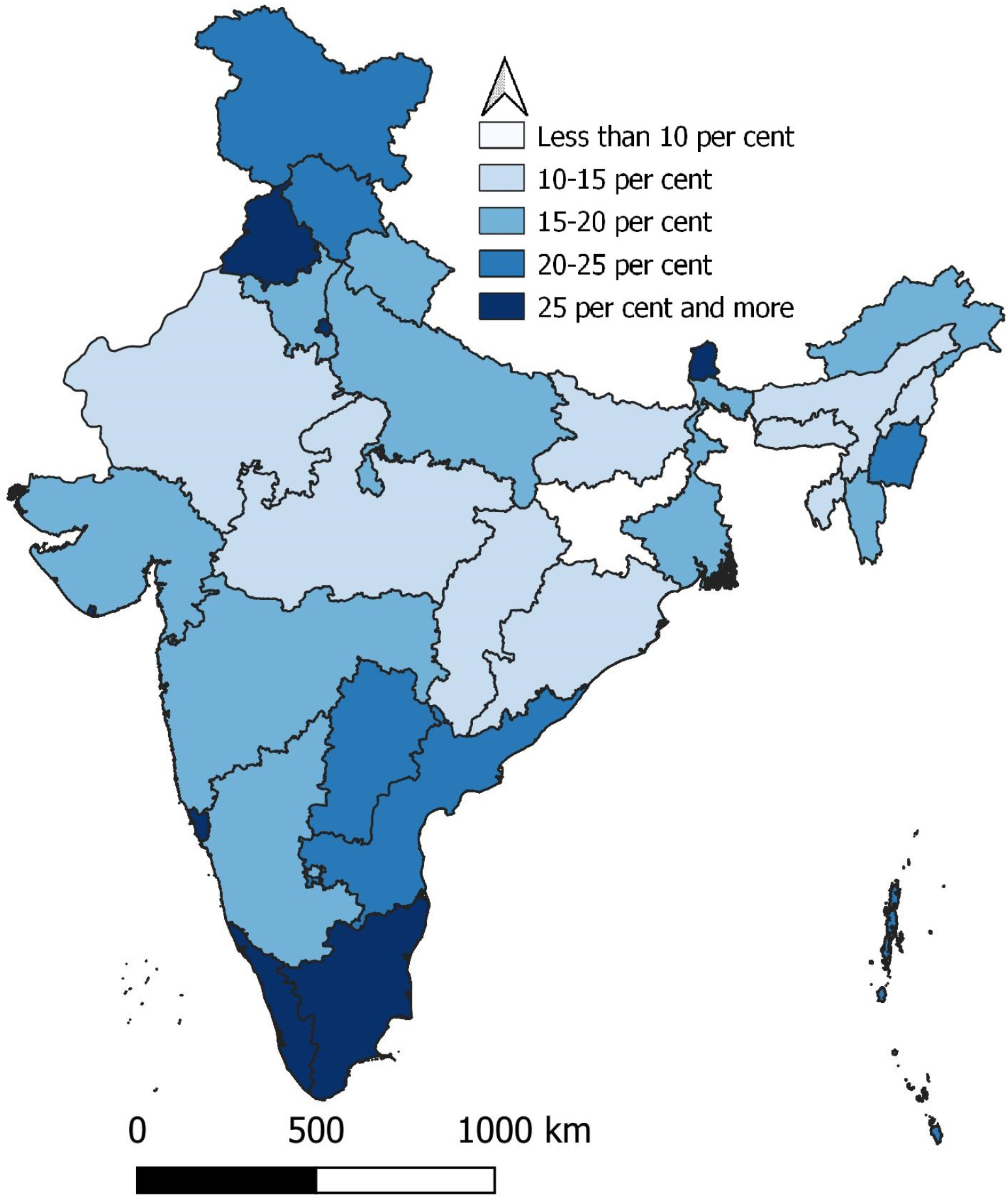
Proportion of adults aged 20-49 years pre-obese 25≤BMI<30.

**Figure 5.**
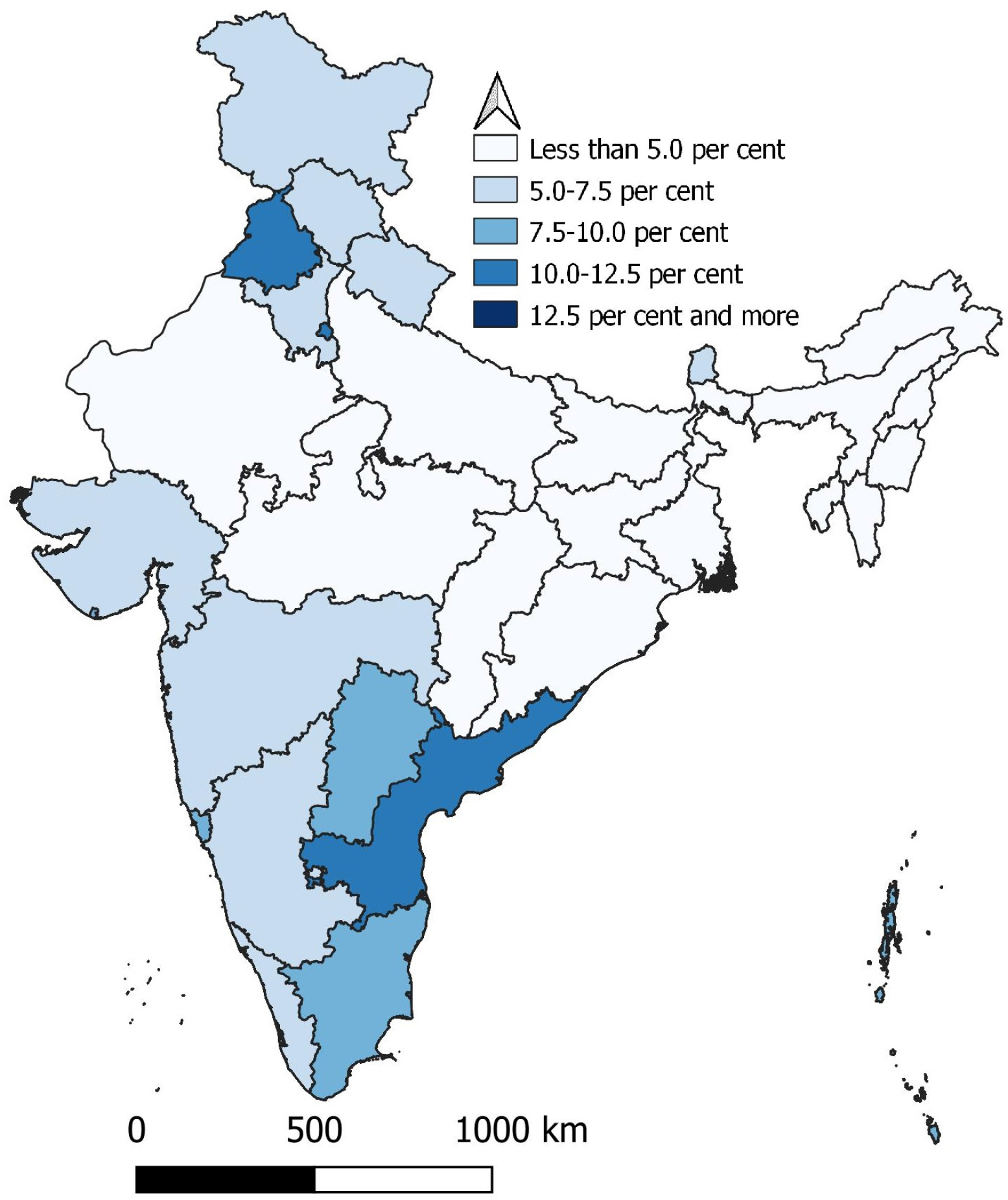
Proportion of adults aged 20-49 obese BMI≥30.

**Figure 6.**
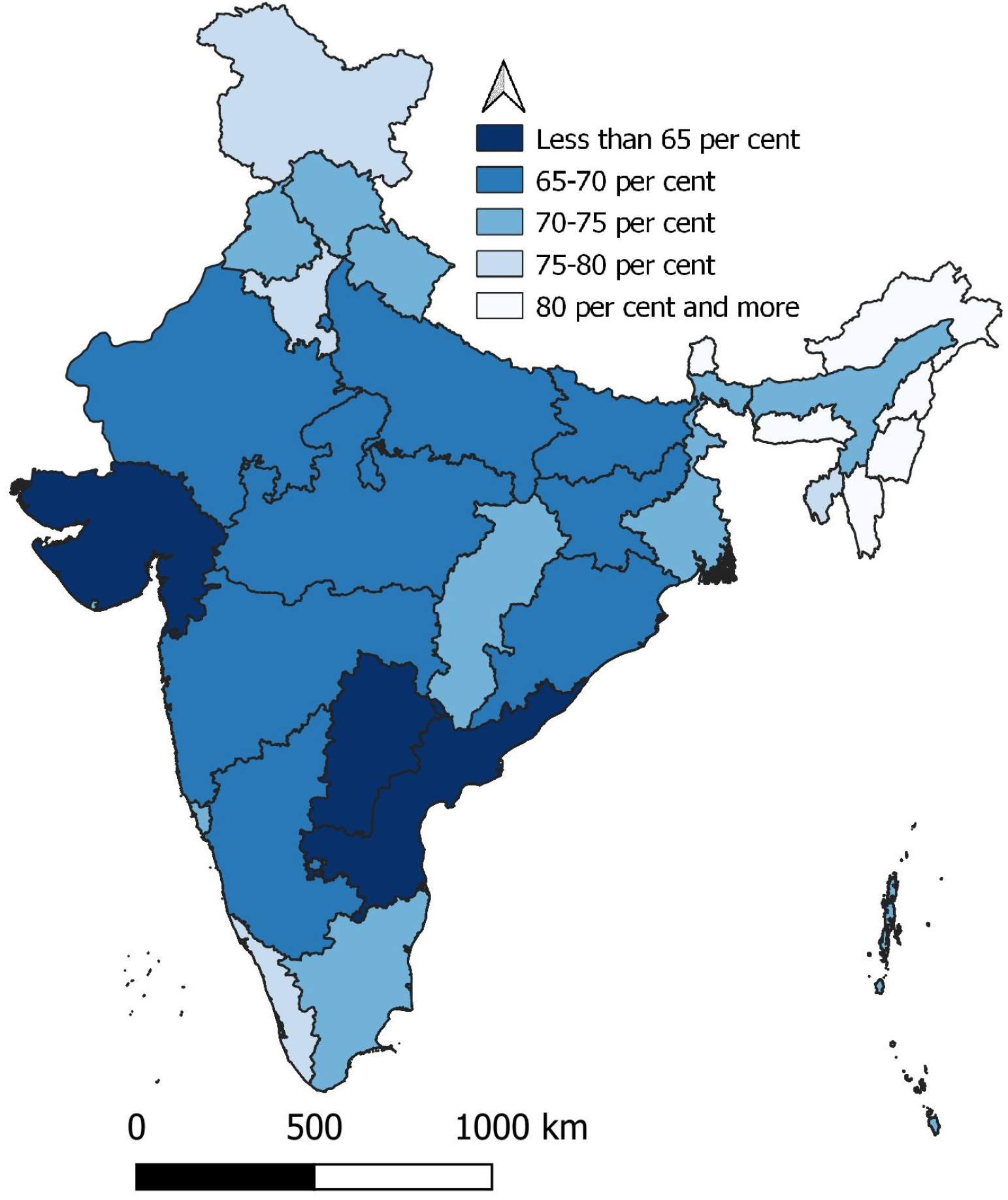
Proportion of adults aged 20-49 years at low to medium risk of type 2 diabetes and cardiovascular disease.

**Figure 7.**
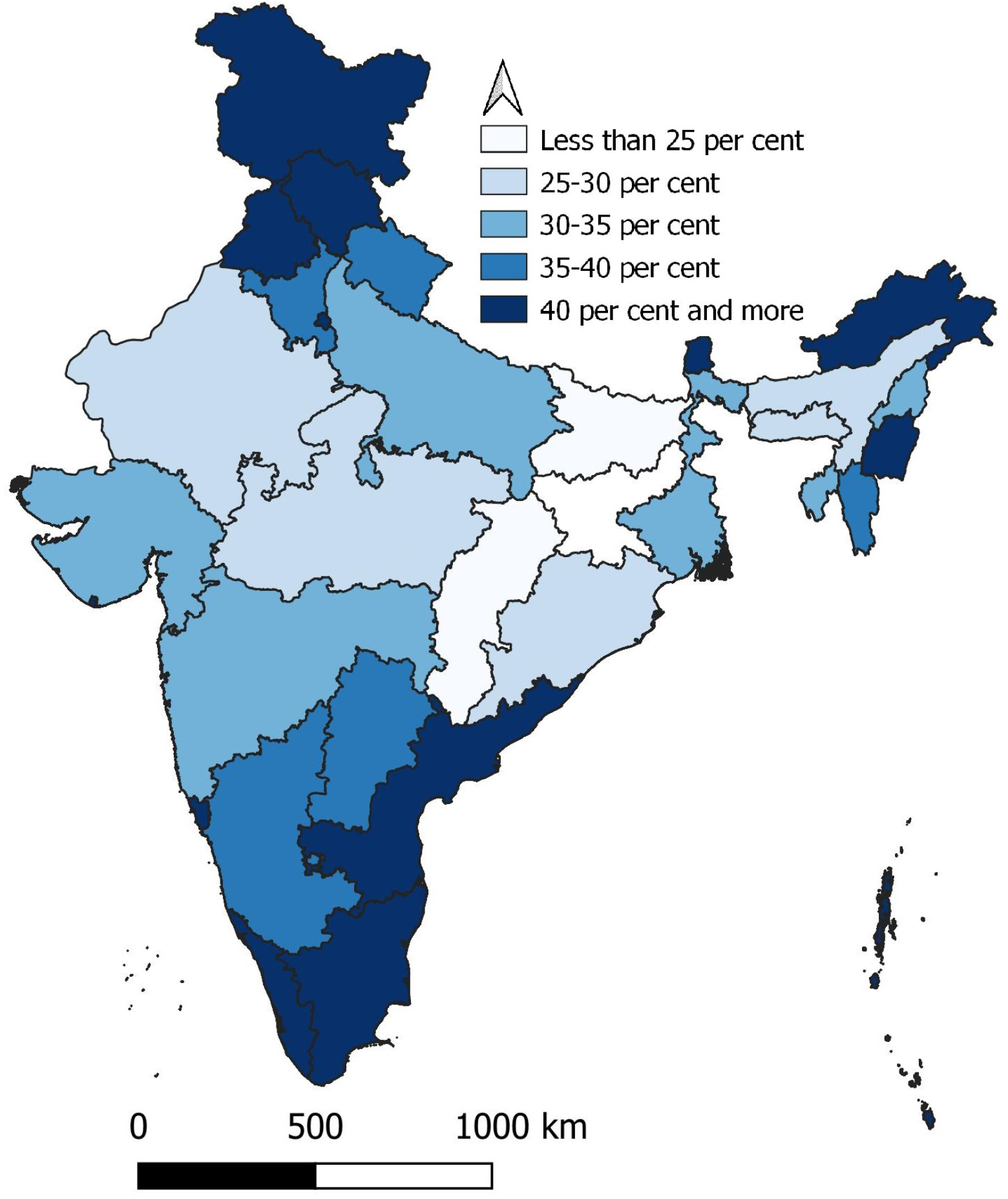
Proportion of adults aged 20-49 years at medium to high risk of type 2 diabetes and cardiovascular disease.

**Figure 8.**
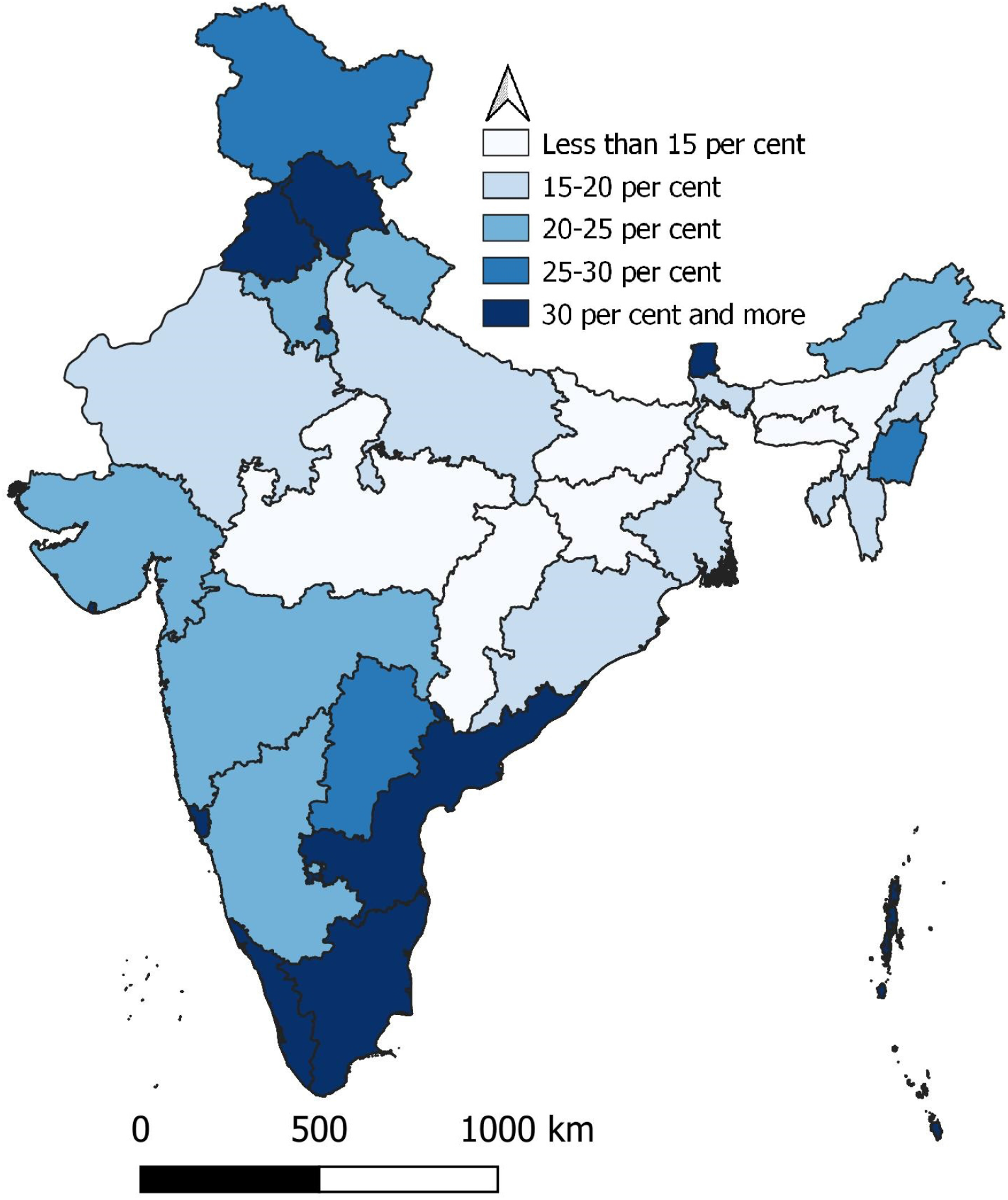
Proportion of adults aged 20-49 years at high to very high risk of type 2 diabetes and cardiovascular disease.

The mean BMI of the individuals belonging to the 15 terminal nodes ranges from a minimum of 19.905 Kg/m^2^ in adults with the poorest standard of living and without toilet in the household to a maximum of 24.996 Kg/m^2^ in adults with richer or richest standard of living, highest education level below higher, living in the urban areas and belonging to Other Castes. The variation in mean BMI across different groups of adults shows that BMI of adults is influenced by the defining individual and household level characteristics of adults. This implies that the body weight related health and mortality risks are essentially different in different adults having distinct defining characteristics. This observation has implications from health policy and health care planning.

The segmentation or classification analysis also reveals the difference in the distribution of BMI by different defining characteristics of adults. In adults with the poor or the poorest standard of living, the residence of the adult is immaterial but in adults with the richer and the richest standard of living, residence matters. Similarly, in adults belonging to Scheduled Castes, Other Backward Classes and Upper Castes and living in either households without any toilet facility or households having an improved toilet facility, rural-urban difference in the distribution of BMI is quite marked. Another important observation is that the type of toilet is immaterial for adults living in urban areas but it matters for adults living in the rural areas.

Table 5 summarises the distribution of BMI in adults aged 20-49 years in different population groups identified through the segmentation analysis. The table suggests that body weight related health and mortality risks are essentially different in different groups of adults aged 20-49 years. In adults with the poor or the poorest standard of living and without toilet in the household, at least 33 per cent are under-weight whereas in adults living in urban areas, more than 10 per cent are obese which indicate that obesity as a health hazard is largely confined to the urban areas of the country. Table 5 also suggests that the proportion of adults aged 20-49 years having normal weight category ranges from less than 50 per cent in adults with richer and richest standard of living, living in the urban areas, educated at the most up to secondary level and belonging to Upper Castes to more than 70 per cent in adults with the poor or the poorest standard of living, having toilet of any type in the household and belonging to Muslim or Other religions.

**Table 5:** Distribution of BMI (kg/m^2^) risk of type 2 Diabetes and cardiovascular disease in different population groups identified through segmentation analysis.

| Terminal node identifier | Proportion (Per cent) of adults aged 20-49 years |  |  |  | Risk of type 2 diabetes and cardiovascular disease in adults aged 20-49 years |  |  |
| --- | --- | --- | --- | --- | --- | --- | --- |
|  | Under weight<br>< 18.5 | Normal<br>18.5-25.0 | Pre-obese<br>25.0-30.0 | Obese<br>≥30.0 | Low to medium | Medium to high | High to very high |
| 8 | 19.65 | 61.57 | 15.24 | 3.54 | 71.94 | 31.78 | 18.51 |
| 11 | 24.25 | 64.02 | 10.14 | 1.60 | 71.31 | 23.48 | 11.57 |
| 12 | 16.03 | 70.51 | 11.50 | 1.95 | 78.97 | 27.49 | 13.28 |
| 14 | 34.61 | 60.56 | 4.19 | 0.64 | 63.77 | 11.98 | 4.66 |
| 15 | 13.28 | 58.37 | 21.91 | 6.44 | 72.65 | 42.28 | 27.83 |
| 18 | 11.46 | 62.34 | 20.91 | 5.28 | 75.94 | 42.09 | 25.80 |
| 20 | 8.82 | 55.74 | 26.41 | 9.03 | 72.22 | 49.78 | 34.69 |
| 21 | 27.17 | 62.38 | 8.89 | 1.56 | 68.76 | 20.92 | 10.27 |
| 22 | 31.29 | 62.81 | 5.22 | 0.69 | 66.74 | 14.85 | 5.81 |
| 23 | 16.13 | 61.73 | 18.00 | 4.15 | 73.81 | 36.42 | 21.83 |
| 24 | 10.24 | 65.54 | 19.33 | 4.89 | 78.48 | 40.81 | 23.88 |
| 25 | 7.11 | 56.44 | 25.93 | 10.52 | 72.73 | 51.01 | 35.31 |
| 26 | 9.64 | 59.02 | 24.05 | 7.29 | 74.53 | 46.04 | 30.66 |
| 27 | 6.42 | 48.60 | 30.41 | 14.57 | 66.72 | 55.21 | 43.30 |
| 28 | 7.65 | 52.64 | 28.19 | 11.52 | 69.94 | 52.55 | 38.66 |
| All | 18.09 | 60.59 | 16.56 | 4.76 | 71.52 | 33.90 | 20.88 |

Source: Author's calculations

It is also obvious that the risk of type 2 diabetes and cardiovascular disease is different in different population groups. In Upper Castes adults with richer and richest standard of living who are living in urban areas and having, at the most, secondary level education, almost 43 per cent are at high to very high risk of type 2 diabetes and cardiovascular disease. By comparison, this proportion is less than 5 per cent in the poorest adults living in households without toilet. In poor adults aged 20-49 per cent living in households without any type of toilet and belonging to Scheduled Tribes, this proportion is just around 6 per cent.

The variation in mean BMI in adults across the states/Union Territories of the country suggests that body weight associated health and mortality risks are essentially different in different states/Union Territories of the country (Table 6). There are only three states/Union Territories where mean BMI in adults aged 20-49 years is estimated to be more than 24 kg/m^2^.

**Table 6.** Mean and standard deviation of BMI of adults aged 20-49 years in states/Union Territories

| SN | State/<br>Union Territory | BMI |  | Proportion (Per cent) of adults<br>aged 20-49 years |  |  |  | Risk of type 2 diabetes<br>and cardiovascular<br>disease in adults aged<br>20-49 years |  |  | N |
| --- | --- | --- | --- | --- | --- | --- | --- | --- | --- | --- | --- |
|  |  | Mean | SD | Under<br>weight | Normal | Pre-obese | Obese | Low to<br>mediu<br>m | Mediu<br>m to<br>high | High to<br>very<br>high |  |
| AN | Andaman and Nicobar Islands | 23.71 | 4.29 | 8.45 | 60.72 | 22.71 | 8.12 | 74.76 | 46.82 | 30.07 | 2734 |
| AP | Andhra Pradesh | 23.64 | 5.01 | 14.64 | 49.87 | 24.83 | 10.66 | 64.96 | 45.16 | 34.36 | 9453 |
| AR | Arunachal Pradesh | 22.95 | 3.37 | 6.04 | 71.59 | 19.11 | 3.26 | 85.42 | 42.32 | 22.11 | 12814 |
| AS | Assam | 21.38 | 3.60 | 21.71 | 63.68 | 12.47 | 2.14 | 72.47 | 27.56 | 14.49 | 25788 |
| BI | Bihar | 21.14 | 3.80 | 25.35 | 60.39 | 11.65 | 2.60 | 68.28 | 24.38 | 14.00 | 37324 |
| CD | Chandigarh | 24.87 | 5.02 | 10.09 | 43.69 | 30.76 | 15.46 | 60.57 | 56.15 | 44.79 | 634 |
| CH | Chhattisgarh | 21.21 | 3.66 | 22.84 | 64.14 | 10.49 | 2.54 | 71.45 | 23.42 | 12.74 | 22770 |
| DA | Dadra and Nagar Haveli | 22.02 | 4.77 | 23.63 | 54.38 | 16.65 | 5.34 | 64.68 | 30.11 | 21.22 | 787 |
| DD | Daman and Diu | 23.83 | 4.54 | 9.53 | 54.10 | 27.05 | 9.32 | 71.48 | 48.28 | 35.60 | 1427 |
| DE | Delhi | 24.44 | 4.77 | 7.41 | 52.66 | 28.08 | 11.85 | 69.80 | 51.90 | 38.60 | 4345 |
| GO | Goa | 23.87 | 4.68 | 10.69 | 53.61 | 26.69 | 9.01 | 71.05 | 50.29 | 34.78 | 2076 |
|  |  | Mean | SD | Under<br>weight | Normal | Pre-obese | Obese | Low to<br>mediu<br>m | Mediu<br>m to<br>high | High to<br>very<br>high |  |
| GU | Gujarat | 22.12 | 4.79 | 24.01 | 52.72 | 16.99 | 6.28 | 63.72 | 33.20 | 22.37 | 22671 |
| HA | Haryana | 22.77 | 3.98 | 11.41 | 65.44 | 18.10 | 5.04 | 77.68 | 39.78 | 22.70 | 20003 |
| HP | Himachal Pradesh | 23.26 | 4.27 | 11.36 | 57.95 | 23.76 | 6.93 | 73.09 | 43.94 | 30.27 | 10046 |
| JA | Jammu and Kashmir | 23.27 | 4.17 | 9.27 | 62.38 | 21.76 | 6.58 | 76.10 | 43.19 | 27.80 | 23045 |
| JH | Jharkhand | 20.85 | 3.66 | 27.48 | 60.88 | 9.41 | 2.23 | 67.29 | 20.72 | 11.42 | 25437 |
| KA | Karnataka | 22.45 | 4.41 | 17.53 | 58.57 | 17.99 | 5.91 | 70.27 | 36.16 | 23.33 | 24481 |
| KE | Kerala | 23.96 | 3.98 | 6.40 | 58.50 | 28.22 | 6.88 | 77.41 | 54.38 | 34.57 | 10777 |
| LA | Lakshadweep | 24.64 | 5.08 | 10.60 | 46.24 | 28.71 | 14.45 | 62.81 | 53.95 | 42.00 | 1038 |
| MA | Madhya Pradesh | 21.32 | 3.96 | 24.33 | 60.49 | 11.86 | 3.31 | 68.39 | 25.31 | 14.84 | 56731 |
| MH | Maharashtra | 22.11 | 4.39 | 21.20 | 56.72 | 16.96 | 5.12 | 67.57 | 33.86 | 21.64 | 27192 |
| MN | Manipur | 23.19 | 3.67 | 7.04 | 66.20 | 21.94 | 4.82 | 80.96 | 44.97 | 26.52 | 12477 |
| MY | Meghalaya | 21.95 | 3.17 | 10.59 | 75.36 | 12.20 | 1.85 | 84.47 | 29.01 | 13.84 | 7573 |
| MZ | Mizoram | 22.55 | 3.40 | 8.16 | 71.94 | 16.88 | 3.02 | 83.68 | 36.73 | 19.73 | 11224 |
|  |  | Mean | SD | Under<br>weight | Normal | Pre-obese | Obese | Low to<br>mediu<br>m | Mediu<br>m to<br>high | High to<br>very<br>high |  |
| NG | Nagaland | 22.20 | 3.30 | 9.62 | 73.32 | 14.58 | 2.48 | 83.70 | 32.60 | 16.95 | 9505 |
| OD | Odisha | 21.56 | 4.08 | 23.82 | 58.09 | 14.40 | 3.69 | 67.81 | 29.09 | 17.84 | 30611 |
| PD | Puducherry | 24.47 | 4.47 | 7.29 | 50.39 | 31.44 | 10.88 | 69.55 | 54.92 | 41.51 | 3842 |
| PU | Punjab | 24.08 | 4.55 | 8.28 | 56.60 | 25.00 | 10.11 | 72.36 | 50.82 | 34.04 | 18603 |
| RA | Rajasthan | 21.60 | 3.98 | 21.78 | 61.87 | 12.66 | 3.69 | 70.42 | 28.14 | 16.01 | 37241 |
| SI | Sikkim | 23.94 | 3.62 | 3.85 | 64.52 | 25.40 | 6.23 | 80.67 | 53.05 | 31.30 | 4961 |
| TA | Tamil Nadu | 23.60 | 4.50 | 11.21 | 55.42 | 25.21 | 8.16 | 71.73 | 47.32 | 32.67 | 27955 |
| TE | Telangana | 22.64 | 4.80 | 20.29 | 51.71 | 20.38 | 7.62 | 64.03 | 37.85 | 27.30 | 6825 |
| TR | Tripura | 21.91 | 3.60 | 17.03 | 65.65 | 14.73 | 2.58 | 76.29 | 33.00 | 17.18 | 4609 |
| UP | Uttar Pradesh | 21.97 | 4.18 | 19.78 | 60.20 | 15.39 | 4.62 | 70.35 | 31.55 | 19.57 | 81759 |
| UT | Uttarakhand | 22.50 | 4.15 | 14.40 | 63.05 | 17.30 | 5.25 | 74.16 | 35.47 | 22.01 | 14838 |
| WB | West Bengal | 21.93 | 3.95 | 19.34 | 61.16 | 16.08 | 3.42 | 71.98 | 33.47 | 19.27 | 16158 |

Source: Author's calculations

**Table 7.** Distribution of adults aged 20-49 years by different population groups identified through segmentation analysis in different states/Union Territories of India

| State/<br>Union<br>Territory | Terminal node identifier of population groups |  |  |  |  |  |  |  |  |  |  |  |  |  |  |
| --- | --- | --- | --- | --- | --- | --- | --- | --- | --- | --- | --- | --- | --- | --- | --- |
|  | 8 | 11 | 12 | 14 | 15 | 18 | 20 | 21 | 22 | 23 | 24 | 25 | 26 | 27 | 28 |
| AN | 4.10 | 3.29 | 3.40 | 1.17 | 5.30 | 4.35 | 15.95 | 8.49 | 1.02 | 14.67 | 26.52 | 2.82 | 1.65 | 4.46 | 2.82 |
| AP | 21.25 | 1.39 | 0.38 | 0.92 | 9.49 | 1.70 | 7.16 | 14.13 | 2.93 | 18.97 | 3.83 | 2.18 | 3.35 | 3.77 | 8.55 |
| AR | 0.46 | 8.15 | 27.12 | 3.41 | 12.21 | 1.76 | 4.50 | 0.76 | 5.03 | 6.41 | 22.57 | 0.09 | 2.48 | 0.21 | 4.83 |
| AS | 0.28 | 29.13 | 20.95 | 3.09 | 6.56 | 1.52 | 2.69 | 5.18 | 2.97 | 16.71 | 5.85 | 0.93 | 0.95 | 1.27 | 1.91 |
| BI | 3.22 | 8.80 | 2.70 | 21.50 | 5.29 | 0.69 | 1.62 | 36.69 | 1.03 | 12.05 | 1.92 | 0.81 | 0.98 | 0.84 | 1.88 |
| CD | 1.10 | 2.37 | 0.00 | 0.32 | 8.36 | 0.32 | 2.84 | 0.79 | 0.00 | 0.63 | 0.00 | 21.14 | 10.09 | 21.61 | 30.44 |
| CH | 8.34 | 8.08 | 0.36 | 13.31 | 6.66 | 1.38 | 3.71 | 18.00 | 15.92 | 8.53 | 0.29 | 2.51 | 3.43 | 2.68 | 6.79 |
| DA | 7.50 | 3.18 | 1.14 | 4.19 | 17.53 | 0.89 | 2.16 | 0.64 | 34.05 | 5.84 | 0.25 | 5.34 | 4.32 | 4.57 | 8.39 |
| DD | 10.51 | 5.05 | 0.49 | 0.00 | 15.07 | 1.26 | 11.91 | 1.68 | 0.21 | 13.17 | 0.42 | 4.70 | 5.26 | 8.13 | 22.14 |
| DE | 1.66 | 1.40 | 0.37 | 0.00 | 17.65 | 0.37 | 0.99 | 0.28 | 0.00 | 0.25 | 0.00 | 15.10 | 8.79 | 22.23 | 30.91 |
| GO | 7.61 | 1.64 | 0.24 | 0.00 | 8.72 | 8.33 | 21.63 | 2.79 | 1.45 | 10.16 | 2.02 | 6.41 | 5.30 | 10.16 | 13.54 |
| GU | 10.22 | 4.72 | 0.27 | 2.71 | 8.80 | 2.17 | 7.71 | 11.36 | 9.73 | 16.89 | 1.54 | 4.14 | 3.03 | 6.59 | 10.11 |
| HA | 4.70 | 2.61 | 0.92 | 0.25 | 6.07 | 6.85 | 26.65 | 3.91 | 0.06 | 17.60 | 2.33 | 5.62 | 4.32 | 7.11 | 10.97 |
| HP | 6.56 | 4.73 | 1.62 | 0.15 | 1.53 | 10.10 | 24.66 | 4.85 | 0.87 | 35.11 | 4.24 | 1.79 | 0.80 | 1.52 | 1.47 |
| JA | 6.48 | 0.91 | 18.26 | 1.27 | 7.86 | 4.75 | 12.15 | 8.44 | 2.55 | 2.26 | 25.42 | 1.54 | 2.19 | 2.16 | 3.76 |
| JH | 8.04 | 3.75 | 2.23 | 22.88 | 7.74 | 0.68 | 1.40 | 26.08 | 10.84 | 4.00 | 1.56 | 1.99 | 2.45 | 2.05 | 4.30 |
| KA | 15.03 | 5.39 | 0.62 | 1.23 | 11.40 | 1.34 | 4.07 | 19.23 | 3.50 | 19.90 | 2.52 | 0.89 | 4.38 | 1.37 | 9.13 |
|  | 8 | 11 | 12 | 14 | 15 | 18 | 20 | 21 | 22 | 23 | 24 | 25 | 26 | 27 | 28 |
| KE | 0.29 | 2.30 | 0.32 | 0.10 | 10.10 | 13.01 | 18.09 | 0.36 | 0.12 | 18.08 | 9.50 | 4.61 | 6.30 | 4.03 | 12.79 |
| LA | 0.00 | 0.00 | 0.19 | 0.00 | 30.15 | 0.87 | 6.17 | 0.00 | 0.00 | 0.00 | 7.03 | 0.00 | 11.85 | 0.19 | 43.55 |
| MA | 8.74 | 4.54 | 0.62 | 11.01 | 8.10 | 1.23 | 5.07 | 23.24 | 10.86 | 8.76 | 0.75 | 2.80 | 3.12 | 3.25 | 7.92 |
| MH | 11.92 | 5.51 | 1.59 | 3.29 | 10.29 | 2.32 | 6.89 | 13.69 | 4.82 | 14.94 | 3.24 | 3.42 | 4.37 | 6.20 | 7.49 |
| MN | 0.09 | 13.22 | 28.68 | 0.39 | 18.11 | 2.55 | 3.10 | 0.36 | 0.49 | 11.16 | 13.74 | 2.20 | 2.35 | 1.73 | 1.83 |
| MY | 0.88 | 2.96 | 34.70 | 0.82 | 12.33 | 1.31 | 2.40 | 0.30 | 4.70 | 1.74 | 30.61 | 0.00 | 3.50 | 0.01 | 3.72 |
| MZ | 0.03 | 0.13 | 15.86 | 0.40 | 18.18 | 1.41 | 7.81 | 0.02 | 0.22 | 0.04 | 26.21 | 0.01 | 6.93 | 0.02 | 22.73 |
| NG | 0.06 | 1.30 | 39.19 | 0.24 | 17.95 | 1.74 | 3.24 | 0.05 | 1.24 | 0.68 | 23.35 | 0.05 | 4.80 | 0.11 | 6.00 |
| OD | 10.00 | 7.77 | 0.82 | 16.22 | 5.13 | 1.20 | 2.55 | 26.23 | 11.30 | 10.61 | 0.44 | 1.75 | 1.44 | 2.14 | 2.41 |
| PD | 11.30 | 0.73 | 0.00 | 0.23 | 17.23 | 2.11 | 4.09 | 7.55 | 0.08 | 5.78 | 0.65 | 0.13 | 18.43 | 0.29 | 31.42 |
| PU | 4.32 | 0.49 | 1.52 | 0.24 | 3.99 | 7.38 | 32.90 | 1.54 | 0.00 | 2.61 | 12.85 | 8.21 | 3.24 | 9.57 | 11.13 |
| RA | 10.86 | 5.27 | 0.67 | 5.21 | 6.37 | 2.60 | 8.71 | 19.38 | 6.49 | 14.31 | 1.51 | 3.99 | 2.96 | 3.98 | 7.67 |
| SI | 0.10 | 3.18 | 3.63 | 0.00 | 14.33 | 2.38 | 5.68 | 0.08 | 0.12 | 27.84 | 29.77 | 1.29 | 2.64 | 3.27 | 5.68 |
| TA | 23.52 | 1.84 | 0.19 | 0.60 | 14.29 | 2.68 | 5.24 | 16.53 | 0.59 | 12.34 | 1.03 | 0.21 | 7.88 | 0.23 | 12.84 |
| TE | 12.29 | 3.63 | 0.66 | 1.55 | 12.37 | 1.26 | 4.48 | 17.48 | 4.15 | 21.47 | 1.80 | 1.74 | 4.26 | 3.24 | 9.61 |
| TR | 0.07 | 42.70 | 11.26 | 1.58 | 15.14 | 0.54 | 1.43 | 0.37 | 0.61 | 15.60 | 3.08 | 1.17 | 2.32 | 1.00 | 3.12 |
| UP | 8.87 | 4.65 | 2.88 | 9.14 | 8.13 | 2.77 | 5.40 | 29.64 | 0.46 | 8.84 | 2.72 | 3.48 | 3.12 | 3.32 | 6.58 |
| UT | 3.94 | 8.46 | 1.21 | 0.58 | 7.79 | 5.48 | 9.08 | 12.17 | 0.62 | 27.20 | 2.42 | 6.13 | 3.05 | 5.71 | 6.15 |
| WB | 2.09 | 18.62 | 9.61 | 7.30 | 12.02 | 0.93 | 2.24 | 16.08 | 3.06 | 14.11 | 4.69 | 1.84 | 1.59 | 2.80 | 3.02 |

Source: Author's calculations

By contrast, Bihar has the lowest mean BMI and in three other states mean BMI is less than 21.5 kg/m^2^. By contrast, in the Union Territories of Chandigarh, Lakshadweep and Puducherry, more than 40 per cent adults aged 20-49 years face high to very risk of type 2 diabetes and cardiovascular disease but this proportion is less than 15 per cent in Assam, Bihar, Chhattisgarh, Jharkhand, Madhya Pradesh and Meghalaya. Obviously, a different strategy needs to be adopted to meeting the health care needs of adults aged 20-49 years in different states/Union Territories of the country.

One reason for the observed variation in the distribution of BMI among adults aged 20-49 years across states/Unition Territories of the country is the variation in the distribution of adults aged 20-49 years in different population groups identified through the segmentation analysis. In the Union Territory of Chandigarh, almost 83 per cent of the adults aged 20-49 years are classified in groups with node ID 25, 27 and 28 and in all these three groups, the mean BMI is more than 24 kg/m^2^. Similarly, in the Union Territory of Lakshadweep, more than 43 per cent of the adults and in the Union Territory of Puducherry, more than 31 per cent of the adults aged 20-49 years are classified in the group with node ID 28. By contrast, almost 37 per cent of the adults aged 20-49 years in Bihar; more than 26 per cent adults aged 20-49 years in Jharkhand; and around 23 per cent adults aged 20-49 years in Madhya Pradesh are classified in group with node ID 21 which has very low mean BMI.

## Discussions and Conclusions

The present analysis presents the pan-India picture of the weight relative to height among adults aged 20-49 years which has implications for health policy and planning. The analysis also shows that the distribution of BMI is different in different groups of adults aged 20-49 years having distinct personal and household level characteristics which essentially means that the health and mortality risks associated with different weight relative to height categories are essentially different for different groups of adults aged 20-49 years. This implies that any approach to meeting the health care needs of adults aged 20-49 years should take into account the variation in weight related health and mortality risks in different groups of adults aged 20-49 years. At the one hand, a large proportion of adults aged 20-49 years belonging to households with poor and the poorest standard of living conditions are under weight. On the other hand, a substantial proportion of adults belonging to households with richer and the richest standard of living are over weight and obese. The health and mortality implications of under weight are essentially different from those of over weight and obesity. There are many epidemiological studies that have analysed the association of over weight or obesity in adults with mortality. However, information about the effect of low weight relative to height on adult health and mortality is sparse. There are some studies that suggest that under weight adults are at increased risk of death mainly due to external causes such as fraility or alcohol or drug use (Roh et al, 2014). Moreover, among under weight adults, smokers may be regarded as vulnerable population from the perspective of health and mortality. Most of these adults belong to the weakest section of the Indian society - the poorest and not having the basic amenity of the toilet in the household. A low weight for height also compromises the productive cpacity of the adult which results in low income and impoverishment which, in turn, contributes to low weight relative to height. At the same time, a sizeable proportion of riche and the richest Indian adults are over weight which also has implications for productivity and health care. At the same time, both under weight and over weight adults aged 20-49 years in the country are spatially distributed which introduces regional dimension of health and mortality issues related to weight relative to height or the nutritional status of Indian adults. It is, therefore, imperative that the regional dimension of nutrition related health and mortality risk in adults aged 20-49 years are taken into consideration while planning for meeting the health care needs of adults.

From the policy and programme perspective, weight related health issues of Indian adults aged 20-49 years may be divided into health and mortality issues related to under weight and health and mortality issues related to over weight. Both contribute to increasing the overall mortality. Weighing too little can contribute to a weakened immune system, fragile bones and feeling tired which may compromise, often quite substantially, the productivity of the adult. Under weight may also be a contributing factor to mental and neurological diseases. The under weight adults aged 20-49 years are primarily confined to the poor and the poorest households and a major contributing factor appears to be the absence of the toilet in the household.

On the other hand, health issues related to over-weight are well known from a large number of epidemiological studies. Over-weight adults are at increased risk for many serious diseases and health conditions (Roberts et al, 2003; Kasen et al, 2008; Luppino et al, 2010; National Heart, Lung and Blood Institute, 2013). The increased risk of type 2 diabetes and cardiovascular disease in over-weight adults is well documented. The present analysis suggests that the problem of over-weight in adults aged 20-49 years is primarily confined to the rich and the richest section of the community - adults in urban areas and belonging to the elite social class (Upper Castes).

Given the two contrastingly distinct scenario of weight related health and mortality issues in Indian adults, as reflected through BMI, efforts to address the health care needs of the adults aged 20-49 years require a two-dimensional strategy. The first dimension of this strategy must focus on screening and counselling under weight adults and improving their living standards by identifying risk factors for external causes of death, many of which may be modified through appropriate social and behaviour change interventions such as creating toilet facility in every household and preventing smoking, alcohol and drug use. The second dimension of the strategy, on the other hand, must be directed towards addressing the challenge of obesity in the affluent sections of the society primarily in the urban areas. This two-dimensional strategy needs to be evolved through a bubble-up rather than a top down approach of the evolution of the health policy and planning for the health care of Indian adults as the relative importance and relevance of the two dimensions of the strategy is regionally sensitive. India’s latest National Health Policy 2017, however, is silent about the relative weight related health and mortality issues of adults as well as their regional perspective (Government of India, 2017). This is so when an increasing proportion of the Indian population is getting concentrated in the adult age group because of demographic transition and the productivity of the labour force is the prime driver of economic growth in the country (Chaurasia, 2019).

## Data Availability

Data used in the analysis are freely available through the Demographic and Health Survey (DHS) Program.

## Notes

### Competing Interest Statement

The authors have declared no competing interest.

### Funding Statement

There is no funding support.

### Author Declarations

The oversight body of the Institution has approved the study.

